# Impact of YAQ001 on the gut microbiome, permeability and markers of systemic inflammation in patients with cirrhosis

**DOI:** 10.64898/2025.12.01.25341286

**Authors:** Jinxia Liu, Fernando Garcia-Guevara, Jane Macnaughtan, Yi Jin, Ainhoa Ferret, Frederick Clasen, Nicholas Yuen, Ramón Garcia Maset, Javier Ramón-Azcón, Annarein JC Kerbert, Theo Portlock, Javier Martinez, Abeba Habtesion, Alexandra Phillips, Francesco De Chiara, Ganesh Ingavle, Tom Baldwin, Cesar Jimenez, Giacomo Zaccherini, Katherine Husi, Miguel Rodriguez-Gandia, Paul Cordero-Sanchez, Junpei Soeda, Jude A Oben, Karen Church, Jia V. Li, Aarti Jalan, Adria Juanola, Elsa Sola, Simon Eaton, Carrie Morgan, Thomas Avery, Michal Kowalski, Daniel Green, Amir Gander, Lindsey Ann Edwards, I. Jane Cox, Helena Cortez-Pinto, Reiner Wiest, Francois Durand, Paolo Caraceni, Roberto Elosua, Joan Vila, Marco Pavesi, Vicente Arroyo, Nathan Davies, Rajeshwar P Mookerjee, Victor Vargas, Susan Sandeman, Gautam Mehta, Julian R. Marchesi, Agustin Albillos, Jennifer L. Rohn, Fausto Andreola, Saeed Shoaie, Rajiv Jalan

## Abstract

**Background and aims:** YAQ001 is a novel, highly engineered, non-absorbable, gut-restricted, multiporous, carbon bead adsorbent. Pre-clinical studies and a clinical trial confirmed its therapeutic potential and safety in patients with cirrhosis. The aims of this sub study were to evaluate the effect of YAQ001 on the gut microbiome and its association with biomarkers of gut permeability and systemic inflammation.

**Methods:** In total, 28-patients with cirrhosis were randomized (double-blind) to receive YAQ001 (4g once daily) or Placebo. Gut microbiome and biomarkers of inflammation and gut permeability were assessed at baseline, after 4 and 12-weeks. Changes in biomarkers and relationship with microbiome composition and gene profiling were assessed. The effect of YAQ001 on *Klebsiella pneumoniae* bacterial biofilms was also assessed using an *in vitro* gut epithelial model.

**Results:** Alpha and beta diversity at different time points were unaltered. YAQ001 increased the abundance of bacteria associated with improved gut health such as *Adlercreutzia equolifaciens* (p<0.05), a bacterium commonly depleted in liver disease, and decreased the abundance of bacteria associated with infections and poor outcomes such as *Klebsiella pneumonia* and *Streptococcus mutans* (p<0.05 each). YAQ001 impacted positively on virulence factors such as siderophores, fimbriae structures and lipopolysaccharides that are associated with inflammation and invasion (p<0.05 each). Antibiotic resistance genes, decreased significantly in the YAQ001 group. These changes were associated with reduction in markers of gut permeability and systemic inflammation with no significant effect on bile acid metabolism. YAQ001 prevented *Klebsiella pneumoniae* biofilms *in vitro*.

**Conclusions:** The results show that YAQ001 impacts positively on the composition of the microbiome, significantly reduces its virulence, antibiotic resistance genes and biofilm formation, which is associated with modulation of gut permeability and systemic inflammation.

**Impact and implications:** YAQ001 exerts its therapeutic effect by favorably modulating the gut ecosystem in cirrhosis. It selectively increases beneficial gut bacteria while suppressing pathogenic taxa linked to adverse outcomes. Beyond composition, YAQ001 reduces microbial virulence by limiting the production of invasive toxins and lowers the abundance of antibiotic-resistance genes. *In vitro* models also demonstrated its efficacy in inhibiting biofilm formation by key pathogens. In summary, this multifaceted modulation of the microbiome led to tangible clinical improvements, notably enhanced gut barrier function and reduced systemic inflammation. The data support further development of YAQ001 as a novel microbiome therapeutic.

**Highlights:**

1. In patients with cirrhosis, YAQ001 specifically enhanced beneficial gut bacteria while suppressing pathogenic bacteria linked to adverse clinical outcomes.
2. YAQ001 significantly reduced the genetic expression of critical virulence factors, including siderophores, fimbriae, and lipopolysaccharides, thereby diminishing the potential for microbial invasion and inflammation.
3. YAQ001 administration led to a significant decrease in the abundance of antibiotic resistance genes within the gut microbiome, potentially restoring therapeutic efficacy of antibiotics.
4. *In vitro* studies demonstrated that YAQ001 effectively prevents the formation of *Klebsiella pneumoniae* biofilms, a key mechanism of bacterial persistence and chronic infection.
5. The beneficial microbial shifts were directly associated with clinically relevant improvements in biomarkers of gut permeability and systemic inflammation.

## INTRODUCTION

Cirrhosis pathogenesis has been closely linked to the microbiome, particularly in advanced disease stages.^1–4^ Bacterial products such as endotoxins translocate from the gut to the liver and systemic circulation, driving dysregulated inflammatory responses and subsequent organ injury.^5,6^ Current interventional strategies to modulate this process are limited to the use of oral antibiotics, which aim to reduce bacterial overgrowth and endotoxemia in cirrhosis.^7–9^ Sometimes those treatments necessitate their chronic administration for secondary prophylaxis of hepatic decompensation.^10,11^ While antibiotics may provide short-term benefits, they do not address the underlying gut microbiome dysbiosis and can exacerbate long-term complications, such as the development and accumulation of antimicrobial resistance (AMR) of virulent strains.^12^ AMR has been recognized as one of the greatest challenges facing modern medicine.^13^ Antibiotic administration is particularly common among liver disease patients due to their compromised immune systems. Therefore, there is a critical need for safer, alternative therapeutic strategies that could be used for both primary and secondary prophylaxis, offering a more effective and sustainable approach to managing cirrhosis.

YAQ001 is a novel, highly engineered, non-absorbable, gut-restricted, multiporous, carbon bead adsorbent designed to target gut-related pathophysiological processes.^14,15^ Preclinical studies in cirrhosis models demonstrated its ability to reduce fibrosis, severity of portal hypertension, and organ dysfunction, likely through the restoration of a dysbiotic gut microbiome and reduction in endotoxemia.^16^ A Phase 2, randomized, placebo-controlled trial (CARBALIVE-SAFETY STUDY) assessed the safety and efficacy of oral YAQ001 in patients with decompensated cirrhosis.^17,18^ In this trial, 28 patients with cirrhosis were randomly assigned (double-blind) to receive either YAQ001 (4g once daily) or placebo. Samples, including blood, urine, and stool, were collected at baseline, 4 weeks, and 12 weeks. The primary endpoints focused on safety and tolerability, while secondary endpoints included organ function and nutritional status. No serious adverse events or significant differences in adverse events between groups were observed. Gastrointestinal-related adverse events were noted but were comparable between groups.^16^ Exploratory endpoints investigated microbiome composition and functionality, as well as local and systemic inflammatory indices and markers of intestinal barrier permeability.

This sub study, conducted as part of the CARBALIVE-SAFETY trial, aims to comprehensively explore the effects of YAQ001 on the gut microbiome, including its composition and functional aspects such as metabolism, virulence factors (VFs), antibiotic resistance genes and its effect on bacterial biofilms. We also investigated whether these microbiome changes are associated with changes in markers of gut inflammation, metabolism, and gut permeability, as well as their potential impact on systemic inflammation and bile acid metabolism to better understand the systemic implications of these changes in the context of cirrhosis.

## METHODS

### Study Design and Patients

A group of 28-patients with liver cirrhosis were randomly assigned into Active and Placebo groups as described (NCT03202498) (**Fig.S1**).^16^

Serum and stool samples were collected at three different timepoints: baseline, week 4, week 12. Human stool samples were collected strictly following Strengthening The Organization and Reporting of Microbiome Studies (STORMS) guidelines.^19^ In serum samples, we measured biomarkers associated with gut permeability, systemic inflammation, nutrition, intestine integrity, kidney function, and endothelial function. We also performed targeted metabolomics in serum samples. For fecal samples we measured biomarkers associated with gut inflammation, gut integrity, and metabolism (**Fig.1A**). All the subjects who took at least 70% of the treatment doses (2 sachets/day for Cohort 1) and did not present major protocol non-compliances were included in this analysis. All measures of efficacy including laboratory tests were also performed on this population.

**Fig. 1.**
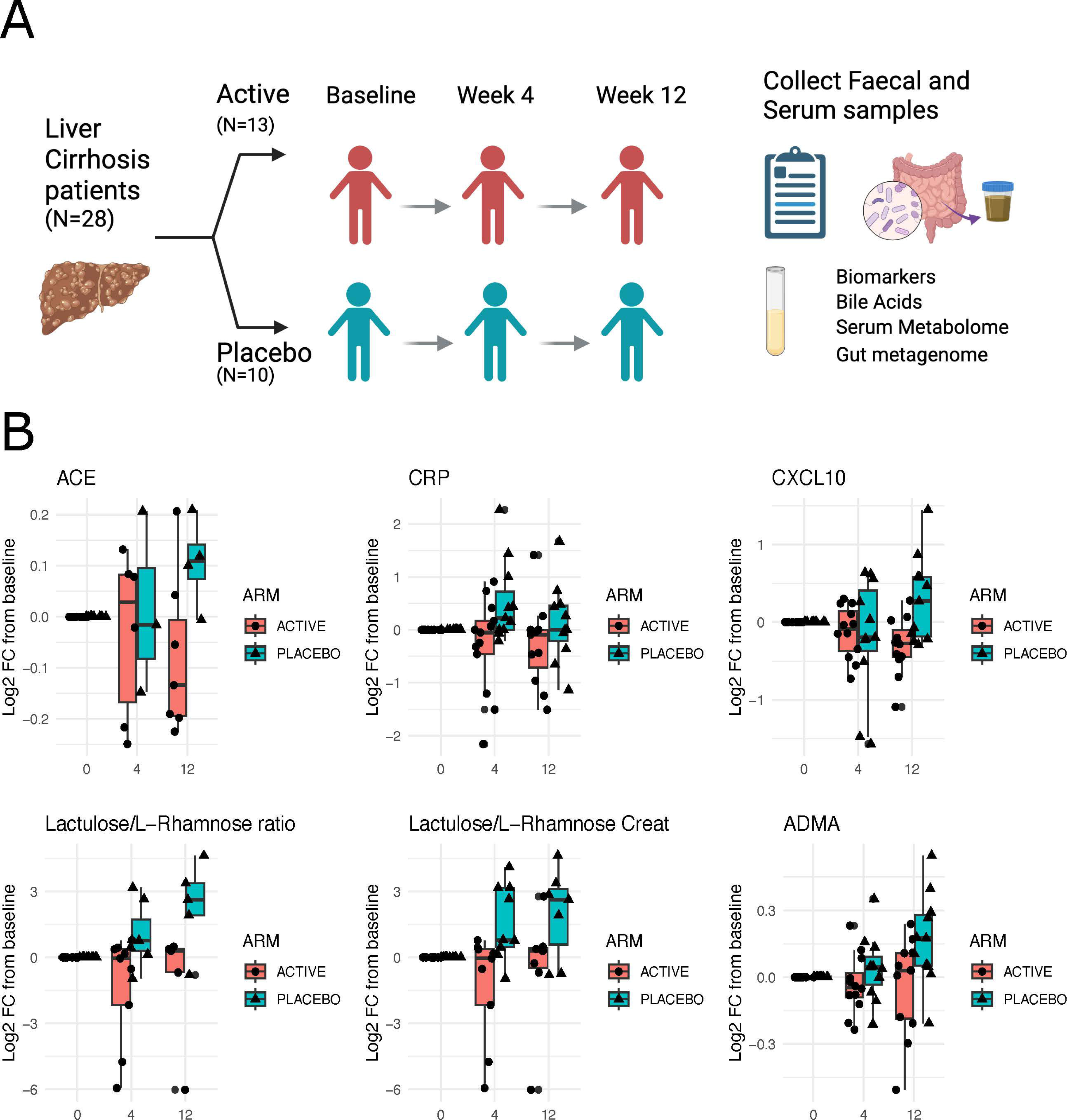
Experimental design and Changes in Biomarkers that changed significantly with YAQ001 treatment. (A) The scheme shows experimental design. (B) Serum biomarkers show improvement in with YAQ001 treatment, the boxplots show the Log2-fold-change in week 4 and week 12 in relation to baseline in Active and Placebo groups. Statistical significance was assessed using two-sided Mann-Whitney U-test and reported as (*) p<0.05.

### Core laboratory analyses

Details of the analysis are available in the Supplementary section.

#### Luminex Assay

The Luminex assays for the measurement of circulating cytokines were performed in accordance with the manufacturer’s instructions. Faecal cytokine analysis and D-lactate assays were performed as previously described.^20^

#### Bile Acids (BAs) Analysis

Cholic acid (CA), chenodeoxycholic acid (CDCA), deoxycholic acid (DCA), lithocholic acid (LCA), ursodeoxycholic acid (UDCA) and their glycine and taurine conjugated species were identified and quantified by ultra-performance liquid chromatography - quadrupole time-of-flight mass spectrometry (UPLC/Q-TOF-MS).

#### Analysis of Short-chain fatty acids (SCFAs) in stool

These were measured using Gas Chromatography-Mass Spectrometry (GC-MS).

### Nuclear Magnetic Resonance (NMR) analysis

Aliquots of plasma (from blood collected into lithium heparin tubes) and urine underwent NMR analyses using standardized protocols.

### Analysis of alterations on the microbiome

#### DNA extraction

For faecal samples, 200 mg of pre-homogenized material was processed using a modified International Human Microbiome Standards (IHMS) protocol.^21^Concentration was measured by Qubit, and quality was verified by 16S ribosomal RNA (rRNA) polymerase chain reaction (PCR) and electrophoresis. Extraction blanks and positive controls were included for quality assurance.

#### Metagenomic sequencing

Stool samples were subjected to metagenomic sequencing using the Illumina HiSeq 2000 platform (Illumina, San Diego, CA), generating paired end reads (2×100 bp). After quality filtering, each sample yielded a minimum of 10 million reads, with an average of 12.5 ± 2.1 million reads (mean ± SD). Only high-quality reads fulfilling these standards were retained for downstream bioinformatic analyses.

#### Gene and Species abundance profiling

The raw reads for all samples were trimmed using Alien Trimmer 0.4.0 with parameters -k 10 -l 45 -m 5 -p 40 -q 20 and Illumina contaminant oligonucleotides.^22^ Trimmed samples were mapped against the Integrated Gut Catalogue 2 (IGC2)^23–25^ using Bowtie integrated into the Metric for Evaluation of Translation with Explicit ORdering (METEOR) pipeline (https://forgemia.inra.fr/metagenopolis/meteor). This generated gene count matrices that were then subjected to downsizing (10 million reads) and normalization (reads per kilo base per million mapped reads (RPKM method)) to generate the gene frequency matrix for downstream analysis. The resulting gene matrices were then projected on previously reconstructed metagenomic species (MGSs)^26,27^ using the top 50 marker genes per MGS to calculate MGS abundances for each sample. Genes included in the IGC2 catalogue are associated with MGSs, we used the taxonomic annotations of the MGSs to assign taxonomic annotations to the genes.

#### Functional annotation of Genes in IGC2 catalogue

Protein sequences of the IGC2 catalogue were annotated for Antibiotic Resistance Genes using the DeepARG software,^28^ using a minimum probability cutoff of 0.9. Proteins of IGC2 catalogue were annotated to Kyoto Encyclopedia of Genes and Genomes (KEGG) orthologs using Diamond against KEGG database^29,30^ Best-hit alignments with e-value < 10^-5^. Annotation of VFs was performed using blastp Best-hit alignments with an e-value cutoff of 1e10^-10^ and a minimum percentage identity of 65% against the set of sequences provided in the virulence factor database (VFDB).^31^ Annotation of Secondary BA genes was performed using best-hit alignments of blastp running against sequences of genes and suggested cutoffs as previously reported (BaiA, HDHA, BaiL, BaiB, BaiCD, BaiH, BaiF, BaiK, BaiG, e-value 1e-10; BaiJ, e-value 1e-5).^32^ To construct the KO, VF, and ARG abundance tables, we started from the gene count table, grouped genes sharing the same annotation (KO, VFID, or ARG), and summed their abundances.

### Analysis of Anti-biofilm and Epithelial Anti-adhesion Activity of YAQ001 against *Klebsiella Pneumoniae* Biofilm formation and treatment assays

Details of the analysis are available in the Supplementary section.

#### Biofilm formation and treatment assays

*K. pneumonia* (TOP52) was pre-cultured statically in TSB at 37 °C for 48 h and seeded into 24-well polystyrene plates (1 mL/well) at an OD_600_ = 0.1. Plates were incubated statically at 37 °C. Total biofilm biomass was quantified by crystal violet (CV) staining. For the biofilm prevention assay, YAQ001 particles (0.625–10 mg/mL in PBS) were added at the time of inoculation, and biofilms were incubated for 72 h. The antibiofilm effect of YAQ001 was investigated against *K. pneumonia* mature biofilms (pre-formed for 48 h) followed by the incubation of YAQ001 particles (0.625–10 mg/mL in PBS) for 24 h. Antibiotic controls were included using both meropenem and ciprofloxacin at 1X to 10000X the Minimum inhibitory concentration (MIC). Antibiotics used in synergy with 10 mM EDTA were also tested. MICs were determined by the broth microdilution method.

#### Polarized intestinal epithelial model and infection

A polarized human intestinal epithelial layer was established by co-culturing Caco2 colorectal epithelial cells and HT29-MTX-E12 mucous-producing, goblet-like colorectal cells at a 3:1 ratio on Snapwell inserts for 14 days. Medium was refreshed every 2–3 days until maturation. For infection, *K. pneumoniae* TOP52 (GFP-tagged)^33^ was grown statically in TSB at 37 °C for 48 h and adjusted to an optical density of OD_600_ of 0.001 or 0.005 corresponding to approximately 1×10^6^ CFU/ml and 2×10^6^ respectively. The apical chamber was infected with *K. pneumoniae* (TOP52) in antibiotic-free medium for 14 h at 37 °C. For treatment, YAQ001 particles (10 mg/mL in PBS) were applied simultaneously with bacteria to a total volume of 500µl. A non-YAQ001 treated control was included for comparison. CFU/ml of planktonic and mucosal associated bacteria were quantified the following day. Confocal microscopy was performed with a Leica SP8 confocal microscope and images were analyzed using LeicaX Software.

### Statistics

Differential abundance analysis of MSP abundance, and genes abundance between Placebo and Active arms was performed using Mann-Whitney U-tests, two tailed, with a cutoff p-value of 0.05, no features were identified as significant after applying False Discovery Rate correction, therefore only p values before correction were employed. No prevalence or abundance filters were applied in abundance tables. For comparing the effect on biomarkers between Active arm and Placebo, first for each subject we measured the Log-2 Fold Change after 4 and 12 weeks in relation to baseline. Then we compared the change between Active and Placebo samples at the same week using a Mann-Whitney U-test. Correlations between MSP abundance and measured biomarkers was performed using Spearman correlation test.

## RESULTS

No significant differences in clinical and biochemical variables, prognostic scores such as the Child-Pugh score [(median (IQR) 7(7–8) vs 7(7–8)] or the Model for End-Stage Liver Disease (MELD) scores [(median (IQR) 13.2 (10.2–16.1) vs 12.6 (9.7–13.5)] were observed between Placebo and Active groups. The rate of adverse events between the two groups were similar. As shown in the previous publication, the nutritional state and associated markers were similar between groups.^16^ Patient details are described in ***Table S1***.

### YAQ001 leads to an improvement in biomarkers associated with systemic inflammation, gut permeability and endothelial function

#### Systemic Inflammation and endothelial function

Of the large number of markers of systemic inflammation measured, we observed significant improvements in the following serum biomarkers in the Active arm in comparison to the Placebo group. Given the clinically stable nature of the studied patients, many of the measurements fell within the normal range and the associated changes in the biomarkers were small. Angiotensin-converting enzyme (ACE) and CXCL10 (**Fig.1B and Fig.S2A, Table S2**) showed a significant decrease after 12 weeks in the YAQ001 and no improvement in the Placebo group. C-reactive protein (CRP) showed an improvement at week 4. ACE is involved in the conversion of angiotensin I into the vasoconstrictor angiotensin II, promoting fibrosis and increased portal pressure within the liver. ^34^ CRP functions as part of the innate immune system by binding to damaged cells and pathogens, rapidly increasing in concentration in the bloodstream when tissue injury or infection occurs. Persistently high CRP levels in cirrhotic patients have been associated with complications such as decompensation, bacterial infections, and mortality. CXCL10 acts as a pro-inflammatory chemokine, attracting immune cells to the site of injury contributing to hepatic inflammation and fibrosis. Asymmetric dimethyl arginine (ADMA) showed improvement at week 12 in the active arm. Increase in ADMA has been associated with impaired liver function. ADMA inhibits endothelial nitric oxide synthase (NOS) activity resulting in endothelial dysfunction and abnormalities of the hepatic microcirculation and portal hypertension. ^35,36^ Additionally, we observed a reduction in cytokeratin-18, M30 in the Placebo group, but the numbers were within the normal range (**Fig.S2F, Table S2**).

#### Gut permeability

We found a significant improvement in the Lactulose/L-Rhamnose ratio, which is a marker of gut permeability (**Fig.1B**).

#### Gut Metabolism

We observed a reduction in the acetic acid, propionic acid, and butyric acid concentrations in the Active arm group (**Fig.S2D**) and no differences in the nutrition biomarkers (**Fig.S2E**). However, the mean value of the concentration at the end of the treatment in the Active arm are 63.5, 7.3, and 14.5 μmol/g wet weight for acetic acid, propionic acid and butyric acid, respectively, which fall within the normal healthy ranges.

#### Markers of gut inflammation

Although there were trends towards lower pro-inflammatory cytokines and endotoxin and, increased anti-inflammatory cytokines in the faeces of the patients in the Active arm, the differences were not statistically different in any of the biomarkers measured in fecal samples (**Fig.S4**).

### YAQ001 modulates the microbiome promoting beneficial species and reducing the abundance of some pathogen associated species

To evaluate the changes in the composition of the gut microbiome to the treatment and placebo, we performed shotgun metagenomics on 70 samples, with average of 15 million reads. The raw sequences were filtered and mapped against gut non-redundant microbial Integrated Gene Catalog 2 (**Methods, Table S3**)^24,25^. After the normalization based on the determined gene counts for each sample, the metagenome species abundances were calculated using the gene counts. After YAQ001 treatment, the species richness between the Placebo and Active groups were similar across all three visits (**Fig.2A**). Additionally, when comparing the distribution of the Bray Curtis distance within individuals at different timepoints between Placebo and Active groups, no significant differences were observed (**Fig.2B**). This could indicate that changes in the overall microbial community composition through time, are similar in both Active and Placebo groups.

**Fig. 2.**
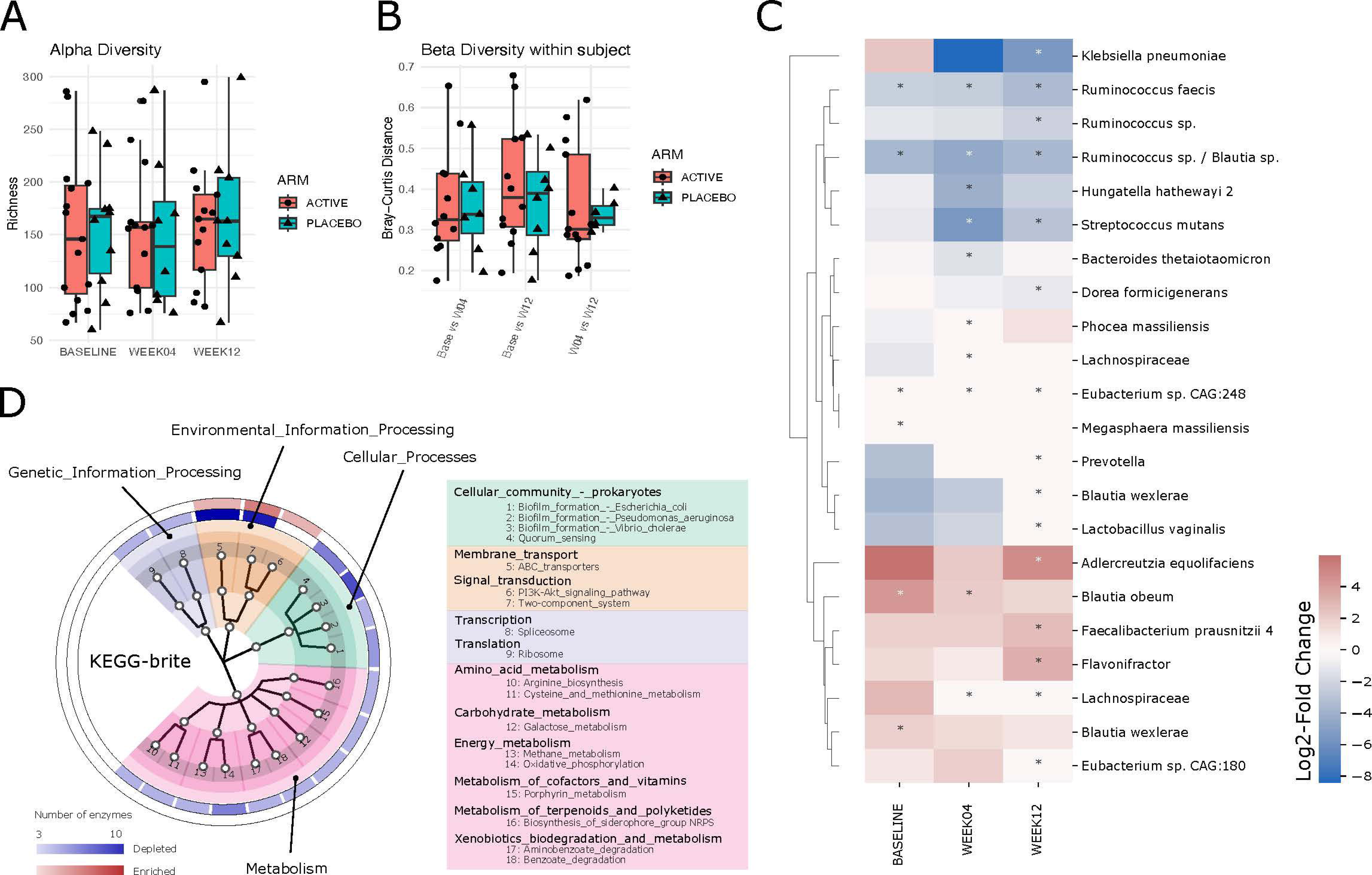
Effect of YAQ001 treatment on microbiome species and functions. (A) Boxplot showing species richness in Active and Placebo arms at baseline, week 4 and week 12. No statistically significant difference was observed between Active and Placebo groups.(B) Boxplots show Beta diversity between time points within the same individual comparing Active vs Placebo groups. No statistically significant difference was observed between active and placebo groups. (C) Differentially abundant species between Active and Placebo Arms, the heatmap shows the log_2_-fold change in mean values of species abundance between Active and Placebo arms. (D) The cladogram shows the KEGG functional categories with at least three genes increasing or decreasing after YAQ001 treatment in both week 4 and week 12, external ring shows the number of genes increasing (red) or decreasing (blue). Statistical significance was assessed using two-sided Mann-Whitney U-test and was reported as (*) p<0.05.

Comparing the effect of YAQ001 with Placebo at the level of individual microbes, most bacteria that increased in abundance in the YAQ001 group have been associated with promoting gut health, such as *Adlercreutzia equolifaciens*, a commensal bacterium with anti-inflammatory properties commonly depleted in liver disease.^37^ Also, we observed increase level of *Faecalibacterium prausnitzii*, a commensal bacteria and significant butyrate producer with anti-inflammatory properties. In contrast, most of the species showing decreased abundance with YAQ001 treatment have been associated with infections and poor outcomes in liver disease patients, such as *Klebsiella pneumoniae*^38,39^ and *Streptococcus mutans^]^*(**Fig.2C, Table S4**).^40^

To reveal the functional landscape of the changes in the gut microbiome, we performed differential abundance analysis of genes grouped by KEGG orthologue (KO) annotations between Active and Placebo arms (**Methods, Table S5**). The functional categories with most genes changing in abundance were the ABC transporters, two-component-systems and biofilm formation genes, all related with external bacterial components. We also found the metabolic pathways of arginine biosynthesis, cysteine and methionine, galactose, methane, oxidative phosphorylation, and biosynthesis of non-ribosomal peptide siderophores had at least three depleted enzymes in the Active arm group (**Fig.2D**). Notably, within the biofilm formation category, only genes decreasing in abundance were observed in the Active arm. Biofilm formation plays a crucial role in pathogenesis, allowing bacteria to adhere to surfaces, providing protection against the immune system and antibiotics, leading to difficult to treat infections. These observations suggest the effect of YAQ001 treatment might impact the membrane or extracellular components of the affected species.

### YAQ001 decreases microbial genes associated virulence and antimicrobial resistance

As observed above, bacterial species decreasing in abundance in the Active arm has been described as pathogenic. Consequently, we performed differential abundance analysis of the microbiome genes coding for virulence factors (VFs) (**Table S6, S7**) comparing Active vs Placebo groups (**Methods**). VFs are bacterial components providing pathogenic bacteria an increased capacity to infect the organism.^41^ All differentially abundant VF genes showed a decrease in abundance in the Active arm compared with the Placebo group, with the greatest decrease at week 12 (**Fig.3A, 3B**).

**Fig. 3.**
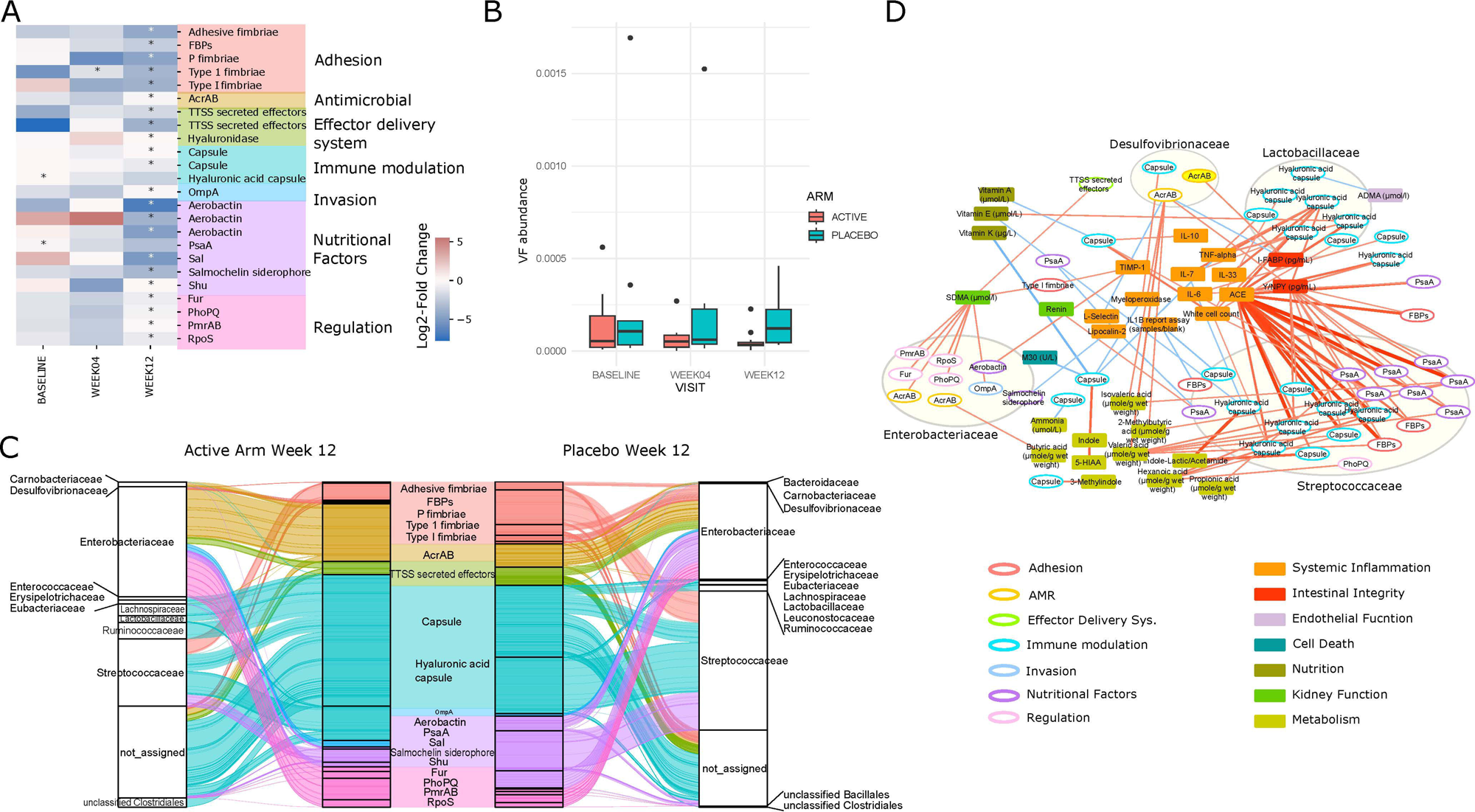
Virulence factors decreasing after YAQ001 treatment. (A) Heatmap showing the log_2_-fold change in mean values virulence factors between Active and Placebo. (B) The boxplots show the sum of the abundance values of all differentially abundant virulence factors in Active and Placebo groups. (C) Alluvial plots show the proportions of the mean abundance values of the virulence factors genes and their associated family of samples at week 12 in the Active (left) and Placebo arm (right). (D) Correlation network between virulence factor genes and measured serum biomarkers: Oval nodes represent genes with virulence factor annotations within a Species. Rectangle nodes represent the measured Serum Biomarkers; Edges represent correlations between a gene and a Biomarkers; positive correlations are shown in red, negative correlations in blue; edge width is proportional to absolute correlation value. Only correlations with p values < 0.002 are shown. Statistical significance in (A) was assessed using two-sided Mann-Whitney U-test and was reported as (*) p<0.05. Correlations in (D) were calculated using Spearman correlation test.

The overall set of decreasing VF genes were linked to adhesion, nutritional factors, effector delivery systems, immune modulation, regulation, invasion, and antimicrobial resistance categories (**Fig.3A, Table S8**). The two most frequent categories were adhesion and nutritional factors. Adhesion VF enable bacteria attachment, invasion, and biofilm formation; for example, fibronectin binding protein (FBP) promotes fibronectin-mediated collagen recruitment, which leads to matrix deposition on and between streptococcal cells to induce the formation of large bacterial aggregates.^42^ In the nutritional VF category, we found mostly genes related to metal ion uptake, like pneumococcal surface adhesin A (PsaA), whose key function is the transport of Mn^2+^ and Zn^2+^ into the cytoplasm of the bacteria; PsaA mutants show marked impact on the capacity to colonize and increased susceptibility to oxidative damage.^43^ Also, within this category, we observed the Sal gene, a major trigger of inflammation and bacterial dissemination during *K. pneumonia* lung infection. Those genes linked to adhesion and nutritional factor categories showed the greatest reduction in the relative abundance at the end of the treatment (**Fig.3C**). A broad range of bacterial families were associated with the changing VFs where the *Streptococcaceae* family showed the greatest changes after treatment (**Fig.3C**).

Interestingly, we identified significant negative correlations between the gene abundance of VFs, like FBP and PsaA, within the *Streptocaccaceae* family and the ACE biomarker, one of the improving biomarkers after YAQ001 treatment (**Fig.3D**).

Identifying VFs as one of the key phenotypic changes and as the YAQ001 treatment reduced some of the opportunistic pathogens, we hypothesized that the treatment could result in a reduction in the AMR genes in the gut microbiome. ARGs pose a significant concern when carried by pathogenic bacterial strains. Moreover, the ARG and VF genes are intertwined and can overlap. Therefore, next we annotated the ARGs using DeepARG software.^28^ Then we investigated the impact of treatment, by performing a differential abundance analysis of predicted ARGs (**Table S9, S10**) between the Active and Placebo groups. Most of the differentially abundant ARGs showed a decrease in abundance in the Active group (**Fig.4A, 4B**). In the multidrug resistance (MDR) family, we observe RamA and OqxB showed the largest (mean log2 fold change <-7), both have an efflux pump mechanism and are also linked to the *Enterobacter* and *Streptococcus* species (**Fig.4A, 4C**). Beta-lactam resistance was another family of genes with dramatic decrease after treatment with significant genes PBP1a and PBP1b linked to the *Streptococcus pneumoniae*. As observed with VFs, ARGs linked to the *Streptococcaceae* and *Enterobacter* family showed the greatest reduction in the Active group (**Fig.4A, 4C**).

**Fig. 4.**
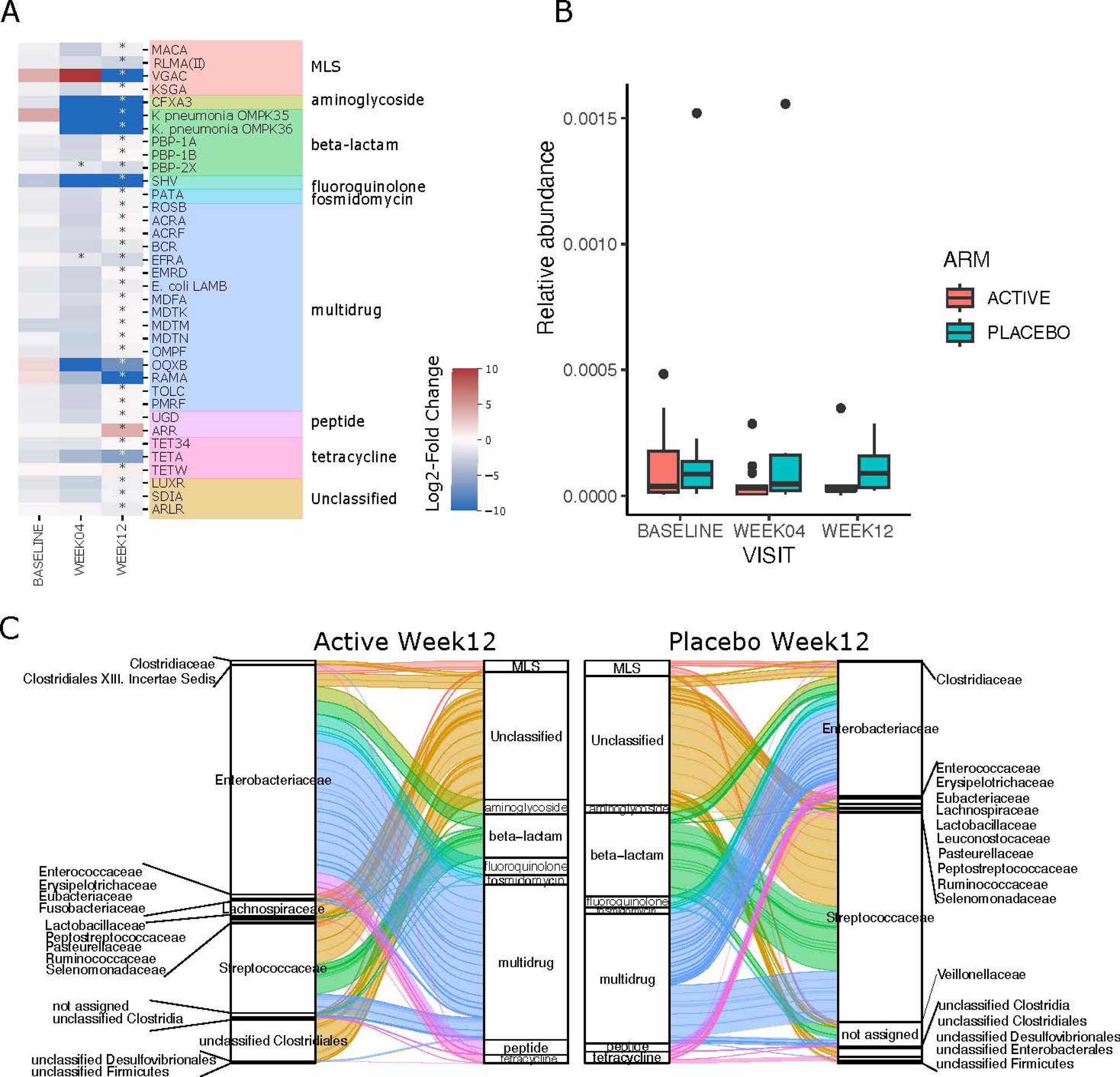
Profile of the antibiotic resistance genes after YAQ001 treatment. (A) Heatmap showing the log2-fold change in mean values predicted ARGs between Active and Placebo. stars (*) p < 0.05 statistical significance was reported as (*) p<0.05. (B) The boxplots show the sum of the abundance values (RPKM) of all differentially abundant ARGs in Active and Placebo groups. (C) Alluvial plots show the proportions of the mean abundance values of the ARGs and their associated family of samples at week 12 in the active (left) and placebo arm (right).

### Effect YAQ001 on fecal and serum BAs and microbiome genes linked to its metabolism

The intestinal microbiota can influence the size and composition of the bile acid pool in serum and alterations in the secondary bile acid pool have been observed in liver cirrhosis patients.^44^ Therefore, we proceeded to test for differential abundance of genes reported to participate in the secondary bile acid metabolism (**Table S11**) and compared these to changes in faecal and serum secondary bile acid concentration (**Table S12, S13**) in relation to baseline in Active and Placebo groups. After YAQ001 treatment, only the BaiCD gene changed showing a decrease in abundance in the Active group (**Fig.S4**). In serum, taurolithocholic acid (TLCA) and glycolithocholic acid (GLCA) showed a trend towards increasing in the active arm but not in placebo at week 12. In fecal samples, glycocholic acid (GCA), taurochenodeoxycholic acid (TCDCA), glycochenodeoxycholic acid (GCDCA), and lithocholic acid (LCA) showed trends to an increase in the Active arm compared to Placebo at week 12 (**Fig.S4**).

### Effect of YAQ001 on the host metabolome

The impact of YAQ001 on metabolic profiles was evaluated through targeted metabolomic analysis at week 4 and 12 relative to baseline (**Fig.S5, Table S14)**. In Active group, significant alterations in metabolite levels were observed compared to the Placebo group. At week plasma tyrosine and phenylalanine showed a marked reduction in the Active arm. These changes persisted through week 12, demonstrating a sustained modulation of metabolic activity in response to YAQ001 treatment. In contrast, the Placebo group exhibited no significant shifts in metabolic profiles at either time point. These results suggest that YAQ001 induces early and persistent modifications in the host metabolome.

### YAQ001 particles prevent biofilm formation of *Klebsiella pneumoniae* TOP52

In order to define the potential mechanisms associated with the selective depletion of pathogenic bacteria such as *K. pneumoniae* with YAQ001, we explored its effect on biofilms generated by this organism in *in vitro* experiments. Crystal violet staining confirmed time-dependent *K. pneumoniae* TOP52 *in vitro* biofilm formation, with maximal biomass detected at 72 h (**Fig.5A, B**). YAQ001 particles did not significantly reduce biofilm biomass against mature biofilms (48h) **(Fig.5D**). Antibiotics, either alone or combined with EDTA, failed to significantly disrupt established biofilms (**Fig.5B**). However, in the biofilm prevention assays, YAQ001 particles reduced biofilm formation in a concentration-dependent manner, with a significant effect at 10 mg/mL (p<0.05) (**Fig.5F**). Meropenem had no biofilm prevention effect at any tested concentration, whereas only ciprofloxacin significantly prevented biofilm formation only at the highest concentrations tested (100x, 1000x, and 10000x MIC). Experimental schematics of antibiofilm and biofilm prevention assays are described accordingly (**Fig.5C, E**).

**Fig. 5.**
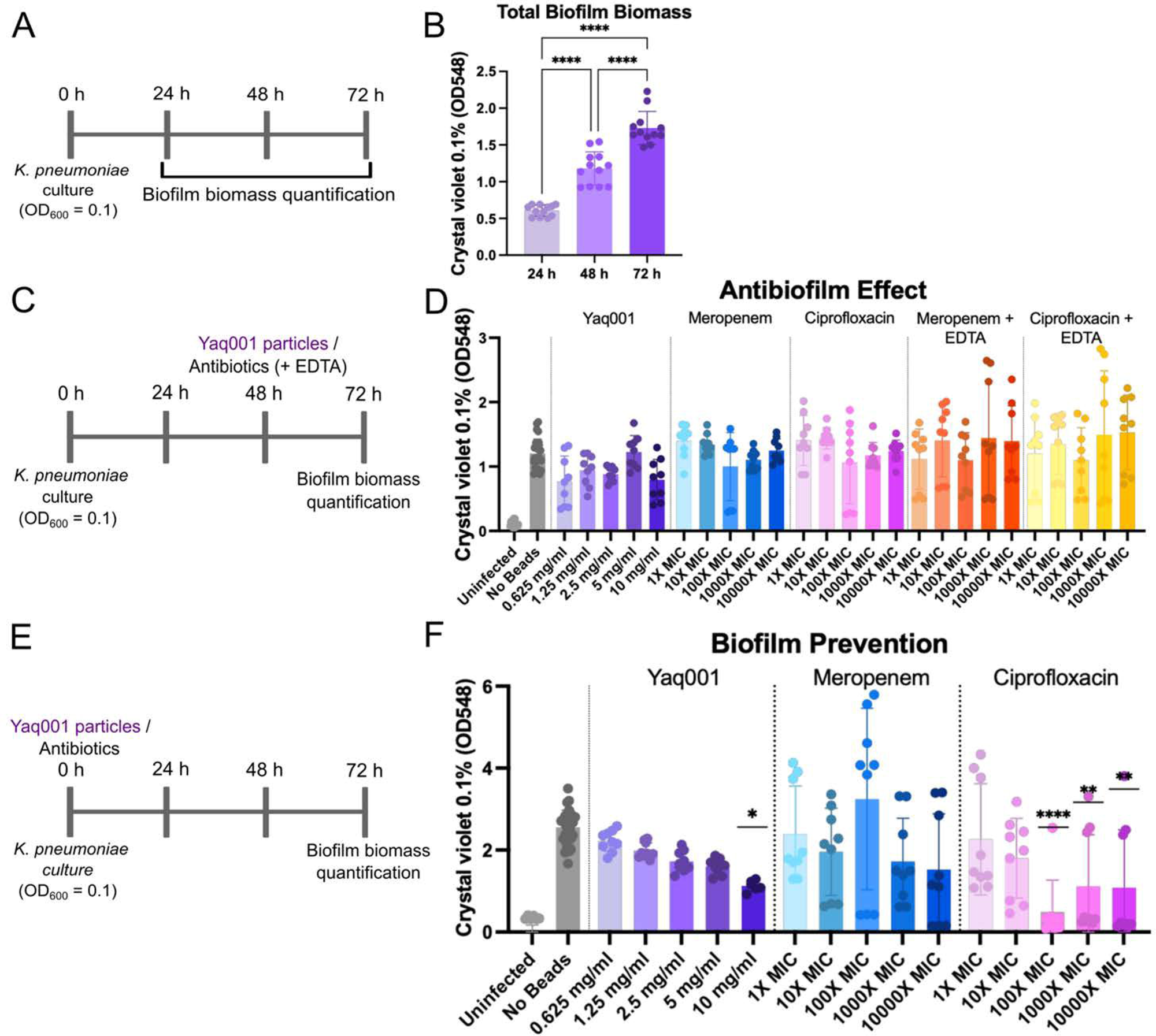
YAQ001 prevents biofilm formation. (A) Biofilm mass quantified at 24-hour, 48-hour and 72-hour time points to determine biofilm maturation. *K. pneum*oniae was cultured in 24-well plates, and YAQ001 or antibiotics were added either at (E) inoculation (prevention) or (C) after 48 h of biofilm formation (antibiofilm assay). (B) Biofilm biomass was quantified by crystal violet staining at indicated time points. (D) Antibiofilm effect of YAQ001 was tested against mature biofilms (48h). (F) Biofilm prevention assay was used to determine the effect of YAQ001 particles to inhibit *K. pneumoniae*. Data represents mean ± SD of three independent experiments. An ordinary one-way ANOVA with Tukey multiple comparisons was used, and statistical significance was reported as (*) p<0.05, (**) p<0.01, (***) p<0.001, (****) p<0.0001.

### *K. pneumoniae* TOP52 binds to the surface of YAQ001 particles and may prevent mucosal/epithelial association in a polarized human intestinal model

To investigate bacterial-epithelial interactions, a polarized intestinal epithelial model was generated using a Caco-2/HT29-MTX-E2 co-culture and infected with *K. pneumoniae* TOP52 with and without YAQ001 particles (10 mg/mL) simultaneously (**Fig.6A**). After 14h treatment, colony-forming units (CFU) of the planktonic fraction did not differ between YAQ001-treated and untreated conditions (**Fig.6B**). In contrast, we observed potentially lower CFUs/ml in the mucosal fraction when treated with YAQ001 compared to the untreated control (data not shown). Via confocal imaging we observed bacterial attachment to both YAQ001 and non YAQ001 treated cells. In YAQ001-treated cells we observed strong bacterial attachment and aggregation of *K. pneumoniae* TOP52 to YAQ001 particles even after multiple PBS washes (**Fig.6D**). The data suggest that the lower mucosal association may be attributed to this mechanism. No bacteria were observed in the uninfected controls (**Fig.6C**).

**Fig. 6.**
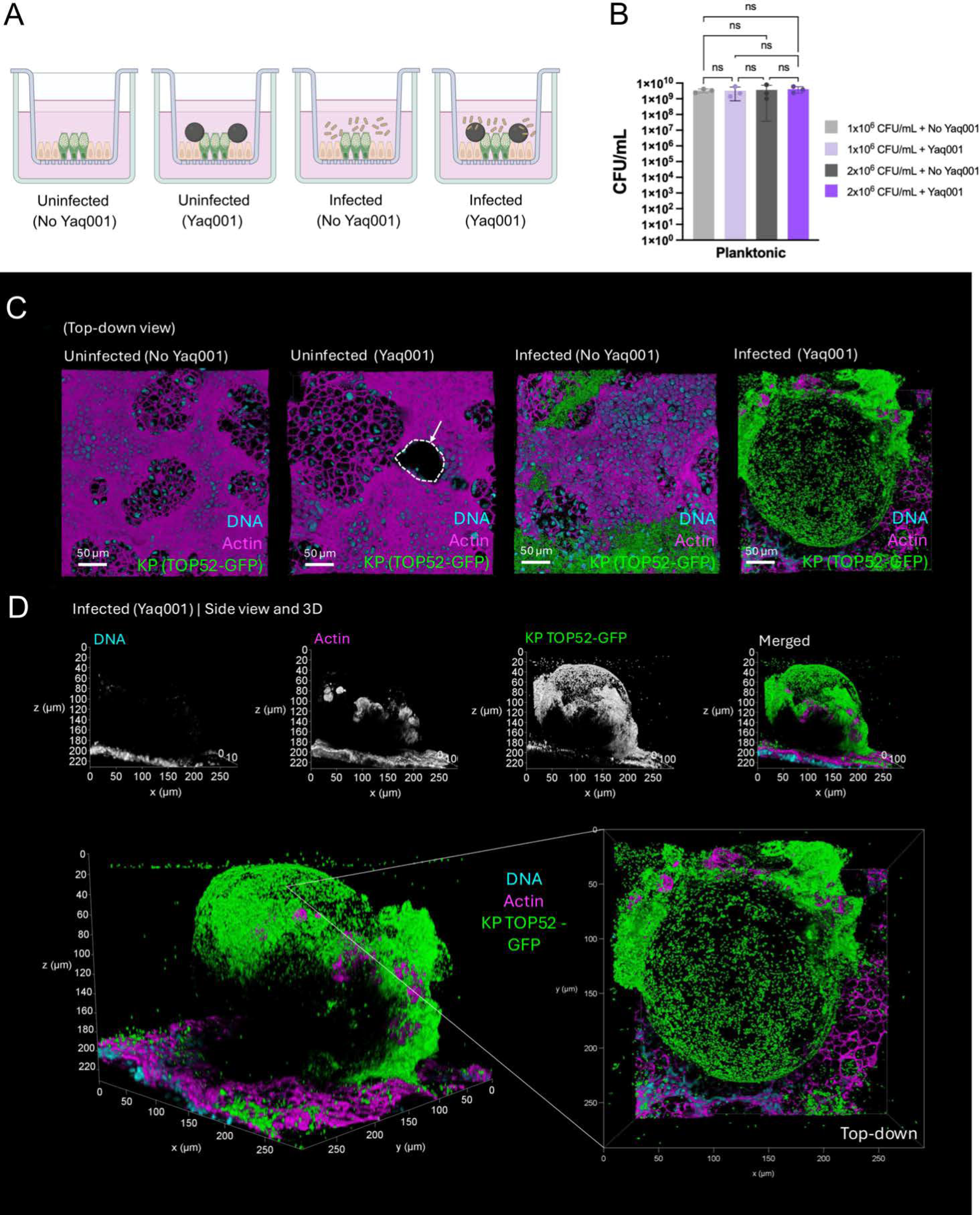
K. *pneumoniae* (TOP52) binds to YAQ001 particles in a human polarized intestinal epithelial infection model. (A) A schematic of polarized intestinal epithelial infection with *K. pneumoniae* TOP52-GFP with YAQ001. Caco-2/HT29-MTX-E2 co-cultures were maintained for 14 days prior to infection with *K. pneumoniae* TOP52-GFP. YAQ001 (10 mg/mL) was applied simultaneously with infection, and outcomes were assessed after 14 h. (B) YAQ001 particles and bacteria were applied to the apical chamber, and planktonic vs. mucosal fractions were collected for CFU analysis. (C) Representative confocal images showing uninfected and infected co-cultures with and without YAQ001 particles. YAQ001 particles are denoted with a white dashed circle. (D) Side view and 3D images indicating bacterial localization with YAQ001. DNA shown in cyan (DAPI), actin shown in magenta (phalloidin-647) and K. *pneumoniae* TOP52-GFP shown in green.

## DISCUSSION

YAQ001 is a novel, orally administered, gut restricted therapeutic approach to sequester substances and metabolites of 1nm to 500nm in the gut lumen by binding through van der Waals forces without exerting any direct antimicrobial activity.^16^ In animal models of cirrhosis, it has been shown to prevent progression of fibrosis, reduction in bacterial translocation, improvement of organ function, occurrence of acute on chronic liver failure with widespread effects on severity of endotoxemia, gene expression in multiple organs and positive effects on the microbiome.^16^ The CARBALIVE-SAFETY clinical trial was a randomized double-blind trial of YAQ001 in patients with cirrhosis, in which its safety and tolerability were confirmed.^16^ In this study, we used the blood, stool and urine samples obtained in the above clinical trial to explore the biological effect of YAQ001 in these patients. The most important biological effect of YAQ001 was observed in the abundance and functionality of the gut microbiome and its impact on gut permeability and some markers of systemic inflammation. We observed a decrease of pathogenic bacterial species and an increase of autochthonous bacteria without major changes in the global microbiome composition. This was associated with functional changes in the microbial genes involved in membrane associated proteins linked to the virulence capabilities, antibiotic resistance and biofilm formation. In *in vitro* studies, YAQ001, prevented the formation of biofilm. These changes in the microbiome were associated with favourable effects on biomarkers representing the intestinal barrier permeability, inflammation and endothelial function.

The metagenomic composition studies showed no significant differences in bacterial diversity, with either YAQ001 treatment or Placebo. This is an important observation as it provides further evidence that YAQ001 does not exert an antibiotic effect and distinguishes it mechanistically from other microbiome targeting treatments such as antibiotics. In a previous *in vitro* study, no effect of YAQ001 on growth characteristics of the bacteria were observed in contrast to antibiotics, which prevented bacterial proliferation.^16^ Shifts in *Klebsiella* and *Streptococcus* in humans compared to favorably with shifts in *Veillonella* and *Roseburia* species in the mouse gut, indicative of a less hostile environment. These data are of particular importance as cirrhosis patients are prone to developing complications such as variceal bleeding, hepatic encephalopathy, ascites, bacterial infections and mortality, which have been associated with gut microbial changes. In cirrhosis patients with progressive disease, pathogenic bacteria such as *Klebsiella* and *Streptococcus* species become more abundant and can cause infections and progression of liver disease.^1,45,46^

As has been shown previously, we observed the presence of multiple microbial VFs, which are known to impact negatively on the outcome of cirrhosis patients.^47^ Although these factors are more pronounced in patients with more severely decompensated disease, it was interesting to note their presence in this population with well compensated cirrhosis. An important observation of this study was the significant impact of YAQ001 on the virulence genes, which encode for multiple virulence factors that extend to mechanisms which allow invasion of the host through secretion of fimbriae, generation of biofilm, secretion of siderophores, attachment processes and secreted lysins. Although the exact mechanism through which YAQ001 exerts this effect is not clear, it may do so by providing a physical barrier selectively decreasing the abundance of virulent species without exerting a selective pressure as is the case with antibiotic administration, which in the long run increases the risk of infection of resistant strains.

AMR is a particular issue in patients with cirrhosis who have greater susceptibility to development of infections due to cirrhosis-associated immune dysfunction.^6,48^ There are several mechanisms that contribute to this immune dysfunction but alterations in the microbiome composition, gut permeability, translocation and inflammation all seem to be important.^6^ ARGs confer microbial communities with the ability to resist antibiotics and represent quorum-sensing and secretion systems that allow microbiota to survive through adaptations in genotypic responses to antimicrobials. The gut microbiome acquires ARGs leading to colonization with pathogenic organisms.^49–53^ The data in our study showed a marked reduction in the abundance of ARGs in the group treated with YAQ001 compared with placebo with significant effects on ARGs associated with several antibiotics including beta lactams, aminoglycosides, MLS and multidrug resistance genes. The data suggest that the reduction in the abundance of the *Streptococcus* and *Klebsiella* species most likely contributes to the reduced burden of ARGs.

The changes in the metagenomic composition due to YAQ001 treatment led us to speculate that some of the differentially abundant extracellular bacterial components, which under ordinary conditions facilitate the binding and invasion might be interacting with the surface of the non-absorbable carbon beads. This property possibly led to a selective decrease of the strains enriched in such elements, along with other genomic elements commonly observed in those strains, such as AMR genes. In turn, this would ameliorate the gut damage inflicted by such bacterial species, and in consequence, lead to an improvement in gut health. To explore the potential operative mechanism through which YAQ001 may alter the composition of the microbiome and its functions, we explored its effect on *Klebsiella pneumoniae* biofilms. The study demonstrated that YAQ001 particles effectively prevented *Klebsiella pneumoniae* biofilm formation *in vitro* at the highest concentration tested. Crystal violet assays indicated concentration-dependent inhibition of biofilm formation by YAQ001, although its effect on pre-formed biofilms was limited and variable—consistent with the known tolerance of mature biofilms to antimicrobials. These findings emphasize the constraints of antibiotic-based treatments in managing biofilm-associated infections and highlight the promise of non-antibiotic alternatives like YAQ001 for biofilm prevention.

The biomarkers related with systemic inflammation showing improvement included ACE, CRP, and CXCL10. Systemic inflammation is central to the pathogenesis of complications of cirrhosis, most markedly acute decompensation and acute on chronic liver failure.^54^ ACE is involved in the conversion of angiotensin I into the vasoconstrictor angiotensin II, promoting fibrosis and increased portal pressure.^55^ We observed strong positive correlations between ACE and virulence factors associated to the species of the *Streptococcaceae* family, suggesting a link between virulence factors and increased ACE levels. CXCL10 is a pro-inflammatory chemokine which has been shown to correlate with acute decompensation, ACLF severity and complications in subjects with severe portal hypertension.^56^ C-reactive protein has been identified as independent predictor of mortality and therefore represents significant target for intervention. Immune cell activation is driven by bacterial translocation in cirrhosis which is a potential mechanism for this observation. These data confirm observations in previous study in animal models.^16^

Endothelial dysfunction is a feature of cirrhosis, which is associated with systemic inflammation.^57,58^ It was therefore not surprising to note a reduction in ADMA in the YAQ001 treated patients, which is commensurate with the reduction in the severity of portal hypertension and improvement of blood pressure and renal dysfunction observed in animal studies. No such correlate was observed in the present study as the patients had relatively compensated cirrhosis.

Existing data reveal that acute decompensation is associated with a highly inflamed and permeable gut barrier.^59^ Decrease in the Lactulose/L-Rhamnose ratio after YAQ001 treatment suggest a stronger gut barrier. Although the exact mechanism has not been elucidated, there were trends towards a positive modulation of fecal cytokines, which are likely contributors. The lack of significant changes in fecal cytokines most likely to reflects the relative normality of the cytokine concentrations, reducing the ability to distinguish any impact of YAQ001.

The data described must be interpreted considering some limitations. The relatively small sample size reduces the power of statistical tests, allowing only the detection of trends towards improvement in several markers but in many cases not reaching statistical significance. In our previous study in animal models, the employed dose was higher, and the animals were more unwell, which may account for more significant effect of YAQ001. Further studies are required to determine an optimal dose. There were trends towards reduction in short chain fatty acids, but the concentrations remained well within the normal range, an observation which should be further assessed, Finally, there was a trend towards higher values of the M30 component of cytokeratin 18, a marker of apoptosis, in the YAQ001 treated patients. As the values were within the normal range, the significance of this observation is difficult to gauge.

In conclusion, these findings support the potential of YAQ001 to address key pathophysiological mechanisms underlying complications of cirrhosis, particularly its effect on the microbiome composition, its virulence and AMR, which were associated with significant impact on markers of inflammation, endothelial function and gut permeability. These results support the development of YAQ001 as a novel prophylactic strategy against biofilm-associated infections, with promising applications in intestinal colonization control and other clinical settings. Larger clinical trials in the prevention of complications of cirrhosis are justified.

## Data Availability

All data produced in the present work are contained in the manuscript.

## Abbreviations

LPS: lipopolysaccharides
AMR: antimicrobial resistance
VF: virulence factors
ARGs: antibiotic resistance genes
STORMS: strengthening the organization and reporting of microbiome studies
PE: Phycoerythrin
Bas: bile acids
CA: Cholic acid
CDCA: chenodeoxycholic acid
DCA: deoxycholic acid
LCA: lithocholic acid
UDCA: ursodeoxycholic acid
UPLC/Q-TOF-MS: ultra-performance liquid chromatography-quadrupole time-of-flight mass spectrometry
SCFAs: short-chain fatty acids
BSA: bovine serum albumin
TBA: tetrabutylammonium
PFBBR: Pentafluorobenzyl bromide
GC-MS: Gas Chromatography-Mass Spectrometry
NMR: Nuclear Magnetic Resonance
PURGE: Presaturation Utilizing Relaxation Gradients and Echoes
PROJECT: Periodic Refocusing of J Evolution by Coherence Transfer
PCA: principal component analyses
IHMS: International Human Microbiome Standards
rRNA: ribosomal RNA
PCR: polymerase chain reaction
IGC2: Integrated Gut Catalogue 2
METEOR: Metric for Evaluation of Translation with Explicit ORdering
MSPs: Metagenomic Species Pan-genomes
MOMR: MetaOMineR
FreqRPKM: frequency Reads Per Kilobase of exon model per Million mapped reads
FPKM: fragments per kilobase of transcript per million fragments mapped
KEGG: Kyoto Encyclopedia of Genes and Genomes
MELD: Model for End-Stage Liver Disease
ACE: Angiotensin-converting enzyme
CRP: C-reactive protein
ADMA: asymmetric dimethyl arginine
NOS: nitric oxide synthase
KO: KEGG orthologue
FBP: fibronectin binding protein
MDR: multidrug resistance
PsaA: Pneumococcal surface adhesin A
TLCA: taurolithocholic acid
GLCA: glycohyocholic acid
GCA: glycocholic acid
TCDCA: taurochenodeoxycholic acid
GCDCA: glycochenodeoxycholic acid
ACLF: acute-on-chronic liver failure.

## Acknowledgements

JRM and the Division of Digestive Diseases receive financial and infrastructure support from the NIHR Imperial Biomedical Research Centre based at Imperial College Healthcare NHS Trust and Imperial College London.

NMR facility at KCL was supported by a funding: The Centre for Biomolecular Spectroscopy, King’s College London is funded by the Wellcome Trust and British Heart Foundation (ref. 202767/Z/16/Z and IG/16/2/32273).

